# Changes in basal insulin secretion capacity after pancreatectomy: a retrospective study

**DOI:** 10.1101/2022.06.14.22276406

**Authors:** Yasuhiro Kihara, Kenta Murotani, Hiroshi Yokomizo

## Abstract

This study evaluated the changes in basal insulin secretion capacity (BISC) after pancreatectomy across two surgical procedures. We also investigated the association between decreased BISC and the introduction of postoperative insulin therapy. The data on 110 patients who underwent pancreatectomy during October 2018–February 2022 at our hospital were analyzed retrospectively. We focused on the C-peptide index (CPI) as an index for BISC. A decreased postoperative BISC was defined as a postoperative CPI (post-CPI) <1.0, which was in turn defined as the event occurrence in our study. The receiver operating characteristic curve for the event occurrence was plotted for factors related to preoperative glucose tolerance. Of the preoperative non-diabetic 73 patients, 44 and 29 who underwent pancreaticoduodenectomy (PD) and distal pancreatectomy (DP), respectively, were examined. A post-CPI of <1.0 was observed in 29 patients (39%). Although the proportion of remnant pancreatic volume was significantly smaller in patients with PD than in patients with DP (38% vs. 68%, p <0.0001), no significant difference was observed in the event rates (39% vs. 41%). In patients with PD, preoperative CPI (area under the curve: 0.75) was used for predicting post-CPI <1.0. Multivariate analysis revealed that preoperative CPI <1.65 (odds ratio: 7.69, 95% confidence interval: 1.87–31.5) was an independent predictor of decreased postoperative BISC. However, in patients with DP, no significant predictors were identified. Induction of insulin therapy was significantly lower in preoperative non-diabetic patients (n=73) after undergoing a pancreatectomy (1.4% vs. 37.5%, p<0.0001) than in preoperative medically treated patients (n=16); there was no significant difference in event (post-CPI <1.0) rates (39.7% vs. 56.2%, p=0.23). Although pancreatectomy reduces BISC after surgery, the coexistence of insulin resistance, which is a pathophysiology of type 2 diabetes mellitus, may play a role in whether postoperative glucose tolerance is reduced to the point where insulin therapy is necessary.

## Introduction

Pancreatectomy is essential in several neoplastic diseases, such as cholangiocarcinoma, cancer of the duodenal papilla, intraductal papillary mucinous tumors, and neuroendocrine tumors, besides pancreatic cancer. Although some postoperative complications, such as pancreatic fistulas, remain to be resolved, surgical techniques and perioperative management have improved the short-term outcomes, such as perioperative mortality [1,2]. Medium-to-long-term complications after pancreatectomy include disease-free survival and deterioration of exocrine and endocrine functions.

Pancreatectomy has an impact on pancreatic endocrine functions; it reduces the capacity for insulin secretion and may lead to the development of new-onset diabetes mellitus (NODM) [3]. Insulin is secreted by β-cells of the islets of Langerhans in the pancreas. Many studies have indicated that the islets of Langerhans are predominantly distributed in the pancreatic tail [4–6]. While some studies suggest that pancreatic tail resection is likely to cause NODM [7], others have reported that the incidence of NODM does not differ between pancreaticoduodenectomy (PD) and distal pancreatectomy (DP) [8,9]. As noted above, some studies examining factors contributing to the occurrence of NODM after pancreatectomy have reported contradictory findings [7–9]. However, only a few studies have evaluated the changes in endocrine function, particularly insulin secretion capacity, before and after pancreatectomy [10,11].

There are two known types of insulin secretion: basal insulin secretion, which remains constant regardless of food intake, and additional insulin secretion, which is in response to rising blood glucose levels post food intake. Decreased basal insulin secretion reduces the blood glucose uptake by the muscles and other tissues and weakens the ability of insulin to inhibit gluconeogenesis in the liver during fasting, leading to increased fasting blood glucose levels, which is a diagnostic criterion for diabetes mellitus (DM). However, the factors affecting changes in the basal insulin secretory capacity (BISC) after pancreatectomy remain unclear. Therefore, in preoperative non-diabetic patients, this study aimed to evaluate changes in BISC after pancreatectomy under two surgical procedures (PD and DP) and identify predictive factors for the postoperative decline in BISC. Further, changes in DM treatment after pancreatectomy were tracked for patients who had received only preoperative oral DM therapy.

## Methods

### Patients

We enrolled 110 patients who underwent pancreatectomy (PD and DP) at Japanese Red Cross Kumamoto Hospital between October 2018 and February 2022. Pre-and postoperative clinical data were retrospectively collected.

Patients were excluded from the analysis if they had undergone central pancreatectomy (n=1), partial resection (n=1), or total pancreatectomy (including total remnant pancreatic resection [n=2]). Moreover, patients with missing data (inadequate preoperative glucose tolerance assessments) were also excluded (n=7). Patients on preoperative insulin therapy were also excluded (n=10). Finally, 89 patients were enrolled (Fig 1). The following main analyses were performed on preoperative non-diabetic patients (n=73), while preoperative type 2 diabetic patients on medication therapy (n=16) were primarily evaluated for the presence of postoperative induction of insulin therapy.

**Fig 1.**
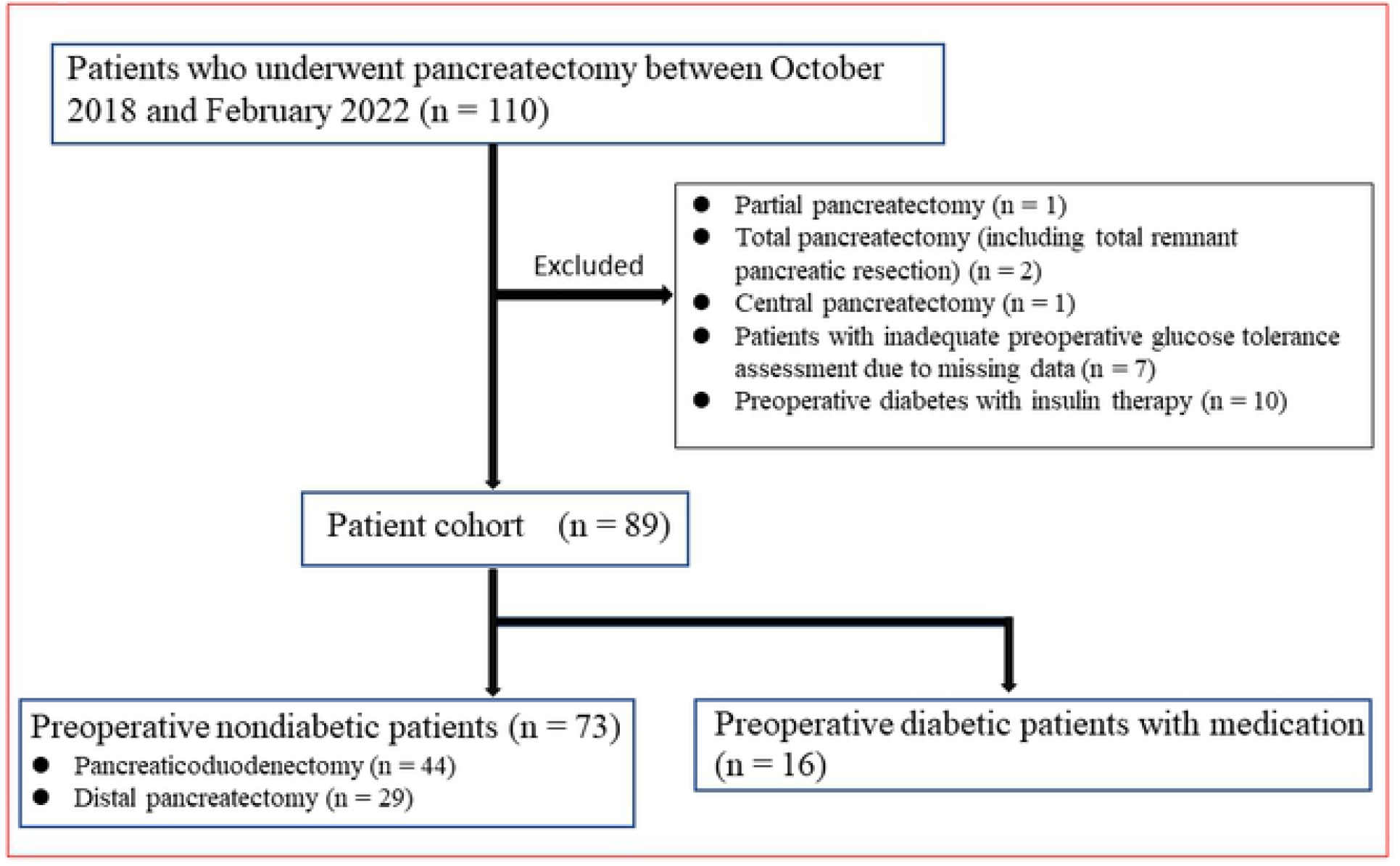
Patient Enrollment Flowchart. CPI, C-peptide index

### Index of basal insulin secretion

Among the various indices of BISC, we used the C-peptide index (CPI), a useful indicator for requirement of insulin therapy [12]. The CPI can be calculated using a simple formula: fasting C-peptide immunoreactivity (ng/mL)/fasting plasma glucose (mg/dL)×100. For normal glucose tolerance, the CPI is approximately 1.60 ± 0.57 [12]. A CPI of <0.8 - 1.0 is indicated as the criterion for insulin therapy requirement for type II DM [12,13]. We defined a decrease in BISC as a postoperative CPI (post-CPI) of <1.0, and the changes in CPI pre-and post-surgery were compared according to the operative procedure.

### Perioperative factors

The surgical procedures of PD and DP were analyzed. Data on operative time, blood loss, blood transfusion, pancreatic firmness in intraoperative findings, postoperative pancreatic fistula (POPF), and delayed gastric emptying (DGE) as postoperative complications were extracted as factors. The POPF was evaluated using the revised classification of the International Study Group of Pancreatic Fistula [14], and DGE was evaluated using the International Study Group of Pancreatic Surgery classification in 2007 [15].

### Preoperative and perioperative factors related to DM and glucose tolerance

We considered patient characteristics, surgery, and DM-related factors as possible predictors of the occurrence of the event of interest (post-CPI <1.0). Patient factors included age, sex, and body mass index; surgical factors included the type of surgical procedure and proportion of remnant pancreatic volume (PRPV) after surgery based on computed tomography (CT) volumetric results; DM factors included preoperative hemoglobin A1c (HbA1c), BISC (CPI), serum insulin level, C-peptide, and index of insulin resistance (Homeostasis Model Assessment of Insulin Resistance).

In Japan, prior to liver surgery, hepatic function is estimated based on the K indocyanine green (K-ICG) value, which is obtained by calculating the extinction rate of intravenously injected ICG. The future remnant hepatic function is estimated by multiplying the K-ICG value by the expected proportion of remnant liver volume [16,17]. However, we introduced a new index, CPV, calculated using the following formula: preoperative CPI (pre-CPI)×PRPV measured by CT volumetry.

Similarly, using the homeostasis model assessment of β-cell function (HOMA-β), that is one index of basal insulin secretion capacity, we generated a new index of HPV, obtained by multiplying preoperative HOMA-β by PRPV, as a predictor of event occurrence (post-CPI <1.0).

Blood tests were performed in the fasting state, and the values were measured preoperatively and within 1 week to 1 month postoperatively. CT volumetry was performed using ZIOstation2 (Ziosoft, Inc., Tokyo, Japan); the pancreatic parenchyma contour was plotted on a 1-mm-width slice from the preoperative and first postoperative CT. The plot area of each slice was calculated, and the pancreatic volume was obtained by integrating the areas.

### Perioperative blood glucose control

In cases of patients with poor preoperative blood glucose control, surgery was performed after inpatient fortified insulin therapy (these patients were excluded in this study). After surgery, a sliding scale was used, and fasting blood glucose level was maintained at <200 mg/dL. When fasting blood glucose level was found to be over 200 mg/dL after food intake was stabilized, oral hypoglycemic agents or self-injection of insulin was introduced under the guidance of diabetes specialists.

### Statistical analysis

To evaluate the predictive power of each factor for event occurrence (post-CPI <1.0), receiver operator characteristic (ROC) curves were generated for each factor, and the area under the ROC curve (AUC) was calculated and compared for each factor. Thereafter, the ROC-AUC variable suggestions found to be the most useful were added to the logistic regression model for event occurrence that was adjusted for age, sex, and surgical procedure to verify whether they remained significant after these adjustments.

Further, we conducted stratified four-fold cross-validation to evaluate the generalizability of the factors selected in the above tests. Specifically, the data were divided into two groups (training and testing groups) at a ratio of 3:1; the data were further distributed such that the proportion of the event occurrence (post-CPI <1.0) was equal between the two groups (training and testing groups). The same division was randomly performed 10 times, and logistic regression was performed to predict the event occurrence (post-CPI <1.0) in the split data sets: using the previously obtained event predictors for each surgery category as explanatory variables—to predict the event occurrence (post-CPI <1.0) for each surgery. Logistic regression was performed using the following formula:

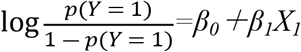

*Y =1*: the event occurrence (post-CPI <1.0), ***β***_***0***_: intercept, ***β***_***1***_: coefficient, *X*_*1*_: obtained predictors of the event occurrence (post-CPI <1.0)

The ROC-AUC was calculated for each split data set and assessed as a mean of the 10 split data sets. Finally, the maximum Youden index (sensitivity-[1-specificity]) was obtained from the ROC curve to calculate the cutoff value of event occurrence predictors for each surgical procedure [18].

Continuous data are expressed as mean ± standard deviation or median and range. Continuous data were analyzed using Student’s t-test or Wilcoxon’s rank sum test for unpaired data and paired t-test or Wilcoxon’s signed-rank test for paired data, as appropriate. Categorical data were analyzed using either the chi-square test or Fisher’s exact test. All statistical analyses were performed using JMP® Pro version 16.0.0 (SAS Institute Inc., Cary, NC, USA) and R version 4.0.0 (The R Foundation for Statistical Computing, Vienna, Austria). All analyses were two-tailed, and p<0.05 was considered statistically significant.

### Ethical statement

The authors are accountable for all aspects of the work in ensuring that questions related to the accuracy or integrity of any part of the work are appropriately investigated and resolved. The study was conducted as per the Declaration of Helsinki (as revised in 2013) guidelines.

The study was approved by the Ethics Board of the Japanese Red Cross Kumamoto Hospital (approval no.: 474), and the requirement for individual consent for this retrospective analysis was waived. We posted a summary of the trial on our website and asked eligible patients to inform us if they wished to be excluded from the study. However, none of the participants requested exclusion.

## Results

Of the 73 preoperative non-diabetic patients included in the analysis, 41 were men and 32 were women (median age, 67.0 years [15–84 years]). The diseases investigated were pancreatic cancer (n=39), biliary tract cancer (n=14), and low malignancy tumor (n=14). The surgical procedures included PD (n=44) and DP (n=29). Four patients who developed NODM postoperatively, one of them was introduced to insulin therapy. The patients’ background characteristics are presented in Table 1.

**Table 1.**
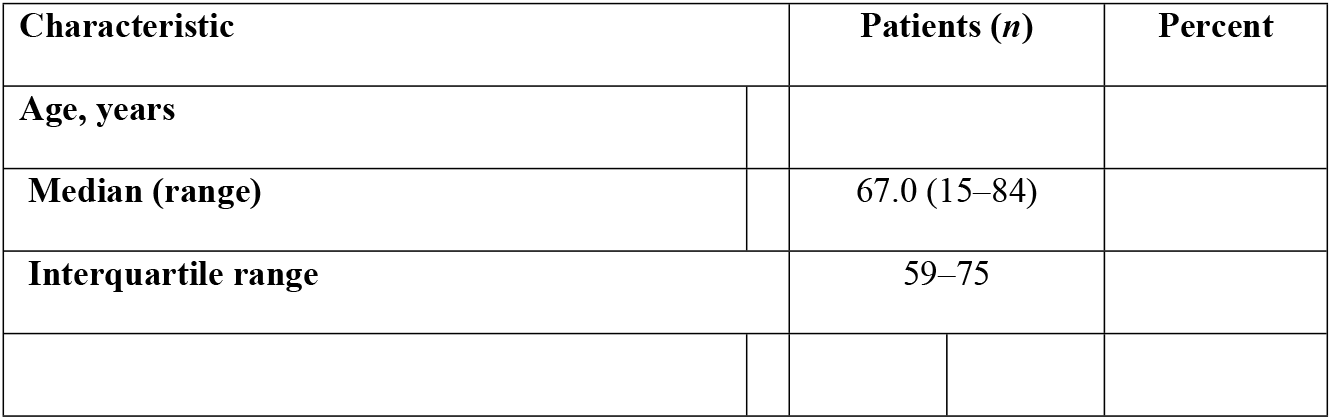
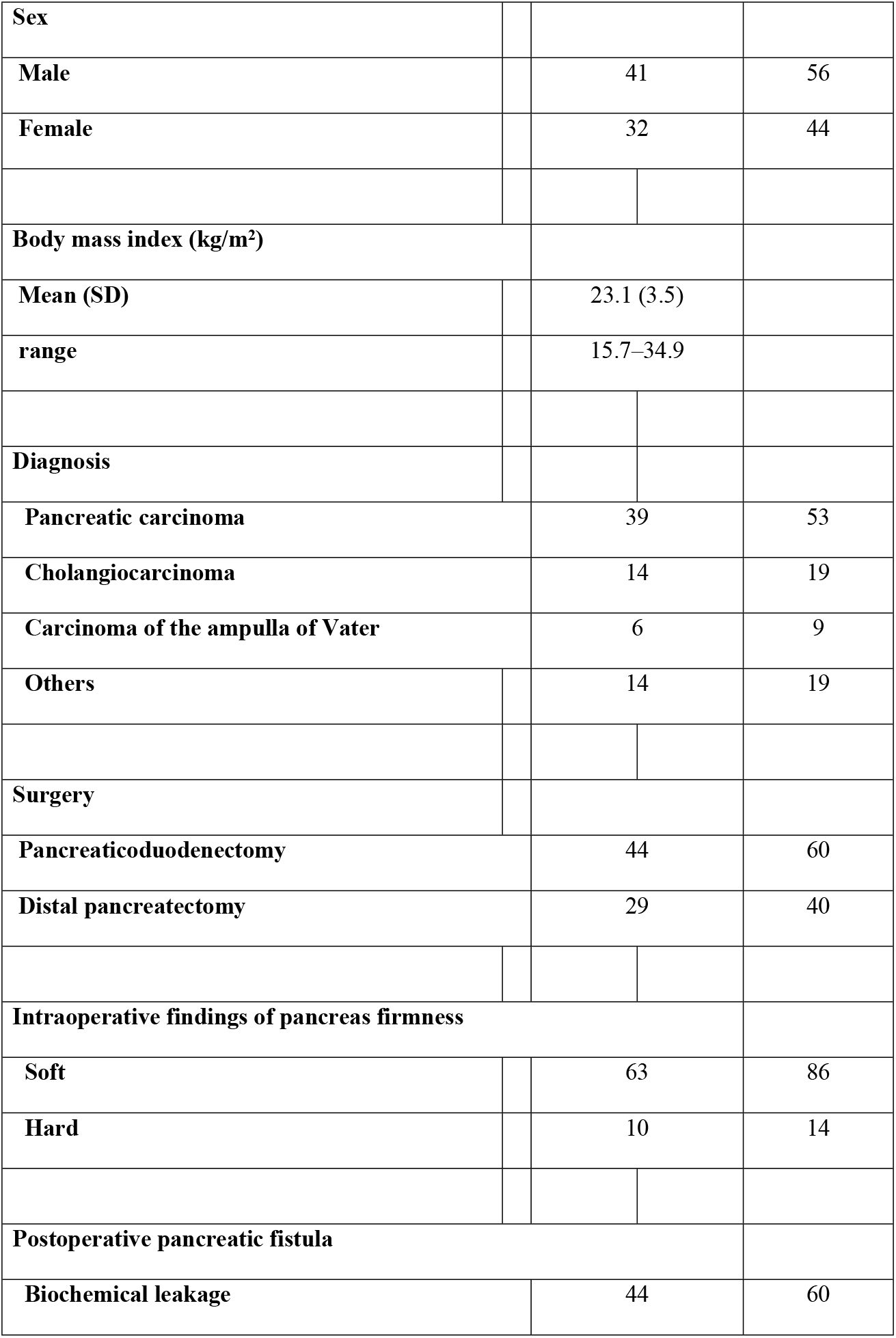
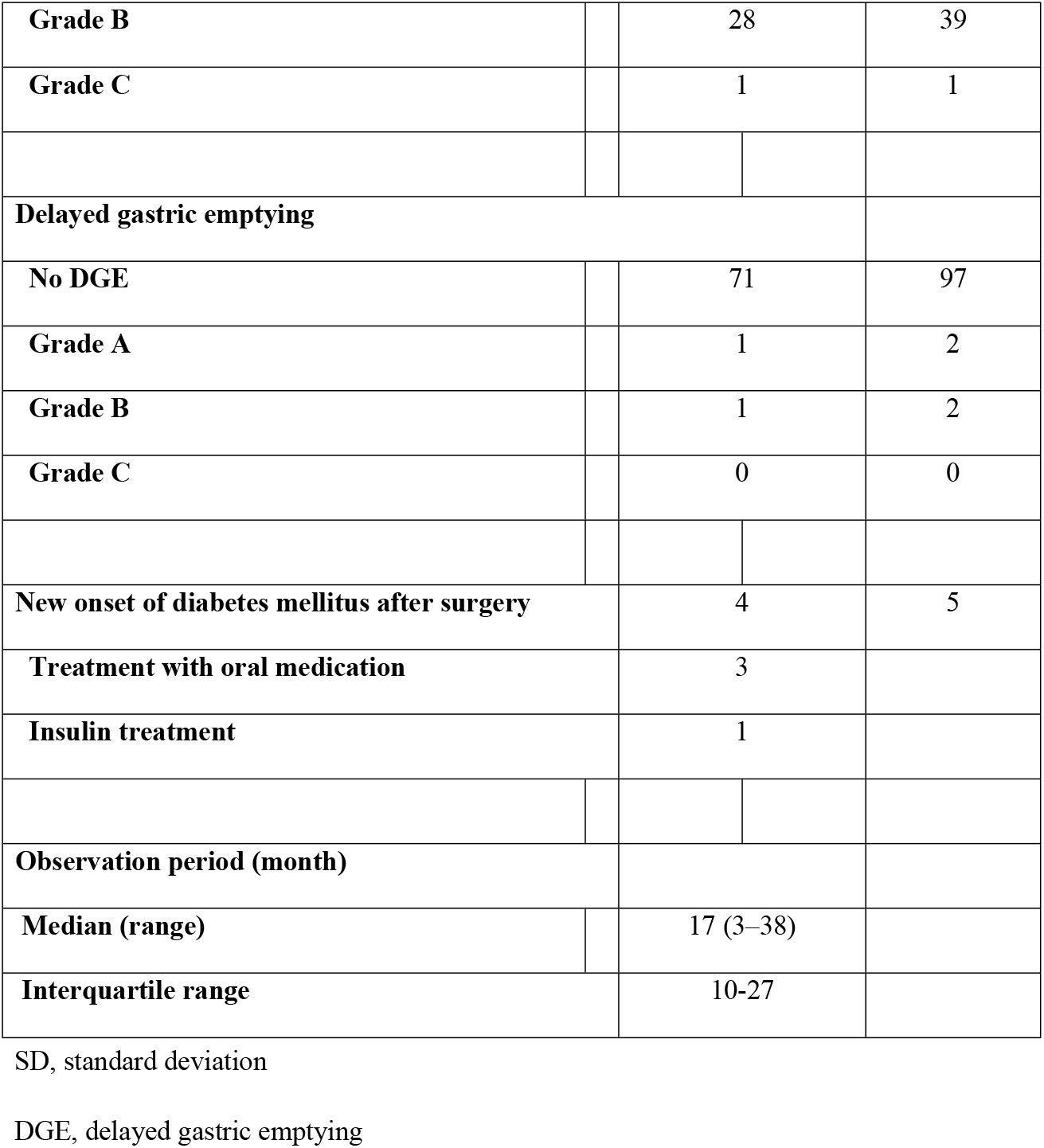
Clinical Characteristics of Preoperative Non-diabetic Patients (n=73).

| Characteristic |  | Patients (*n*) |  | Percent |
| --- | --- | --- | --- | --- |
| Age, years |  |  |  |  |
| Median (range) |  | 67.0 (15–84) |  |  |
| Interquartile range |  | 59–75 |  |  |

|  |  |  |  |
| --- | --- | --- | --- |
| **Sex** |  |  |  |
| **Male** |  | 41 | 56 |
| **Female** |  | 32 | 44 |
| **Body mass index (kg/m<sup>2</sup>)** |  |  |  |
| **Mean (SD)** |  | 23.1 (3.5) |  |
| **range** |  | 15.7–34.9 |  |
| **Diagnosis** |  |  |  |
| **Pancreatic carcinoma** |  | 39 | 53 |
| **Cholangiocarcinoma** |  | 14 | 19 |
| **Carcinoma of the ampulla of Vater** |  | 6 | 9 |
| **Others** |  | 14 | 19 |
| **Surgery** |  |  |  |
| **Pancreaticoduodenectomy** |  | 44 | 60 |
| **Distal pancreatectomy** |  | 29 | 40 |
| **Intraoperative findings of pancreas firmness** |  |  |  |
| **Soft** |  | 63 | 86 |
| **Hard** |  | 10 | 14 |
| **Postoperative pancreatic fistula** |  |  |  |
| **Biochemical leakage** |  | 44 | 60 |
| **Grade B** |  | 28 | 39 |
| **Grade C** |  | 1 | 1 |
| **Delayed gastric emptying** |  |  |  |
| **No DGE** |  | 71 | 97 |
| **Grade A** |  | 1 | 2 |
| **Grade B** |  | 1 | 2 |
| **Grade C** |  | 0 | 0 |
| **New onset of diabetes mellitus after surgery** |  | 4 | 5 |
| **Treatment with oral medication** |  | 3 |  |
| **Insulin treatment** |  | 1 |  |
| **Observation period (month)** |  |  |  |
| **Median (range)** |  | 17 (3–38) |  |
| **Interquartile range** |  | 10-27 |  |

SD, standard deviation
DGE, delayed gastric emptying

Patient factors, surgical factors, and glucose tolerance-related factors were compared according to the operative procedure and the presence/absence of the event (post-CPI <1.0) (Table 2). A post-CPI <1.0 was observed in 29 patients (Table 2, 39.7%). Although PD patients had a significantly smaller PRPV than DP patients (Table 2, PD: 38%, DP: 68%, p<0.0001), no significant difference was noted in the incidence of post-CPI <1.0 (Table 2, 38.6% vs. 41.3%, p=0.81). Comparison of the presence of event (post-CPI <1.0) occurrence revealed significant differences in body mass index (BMI), diagnosis (pancreatic cancer), and glucose tolerance-related factors such as preoperative HbA1c, C-peptide, CPI, HOMA-β, CPV, and HPV (Table 2).

**Table 2.**
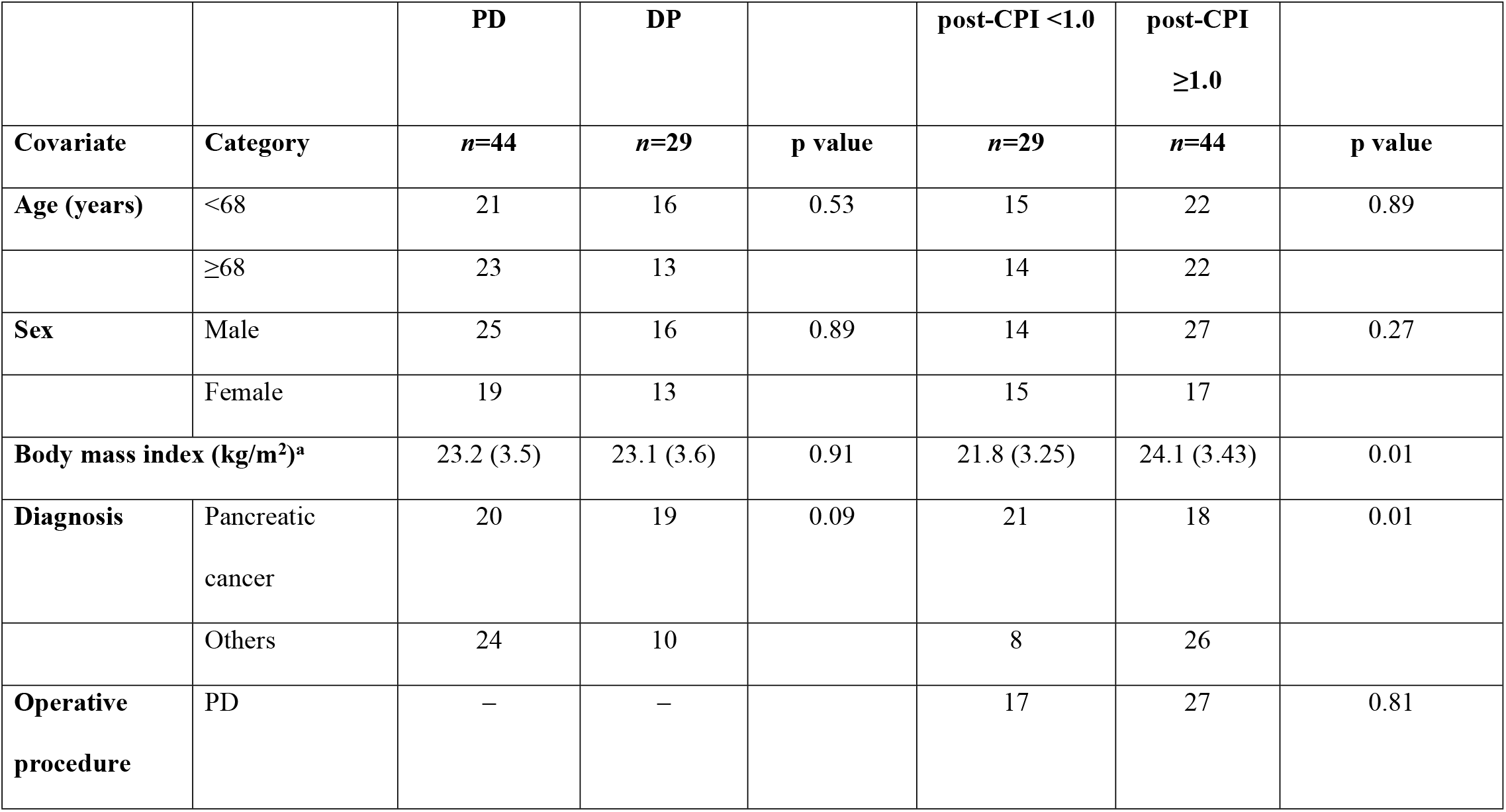
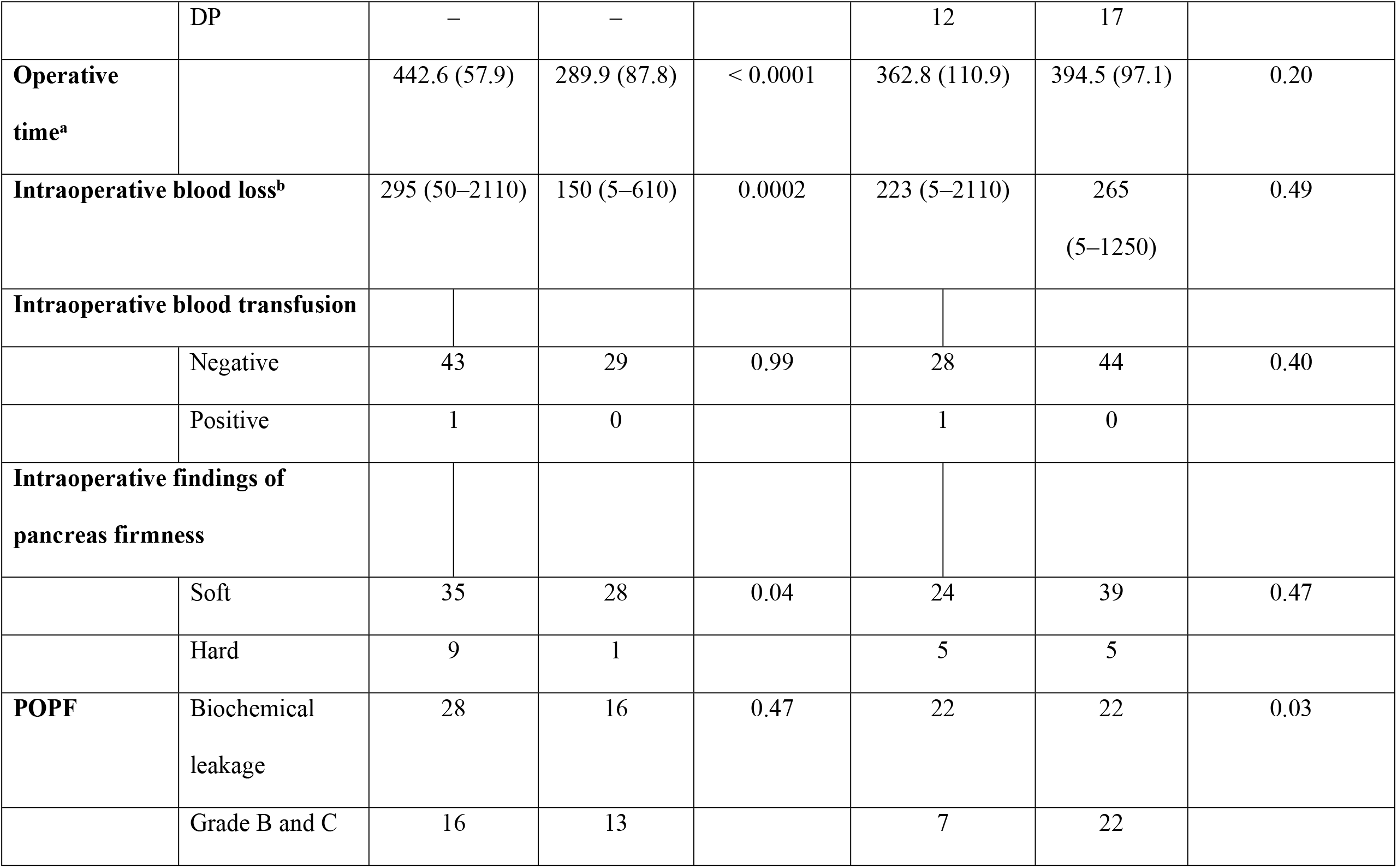
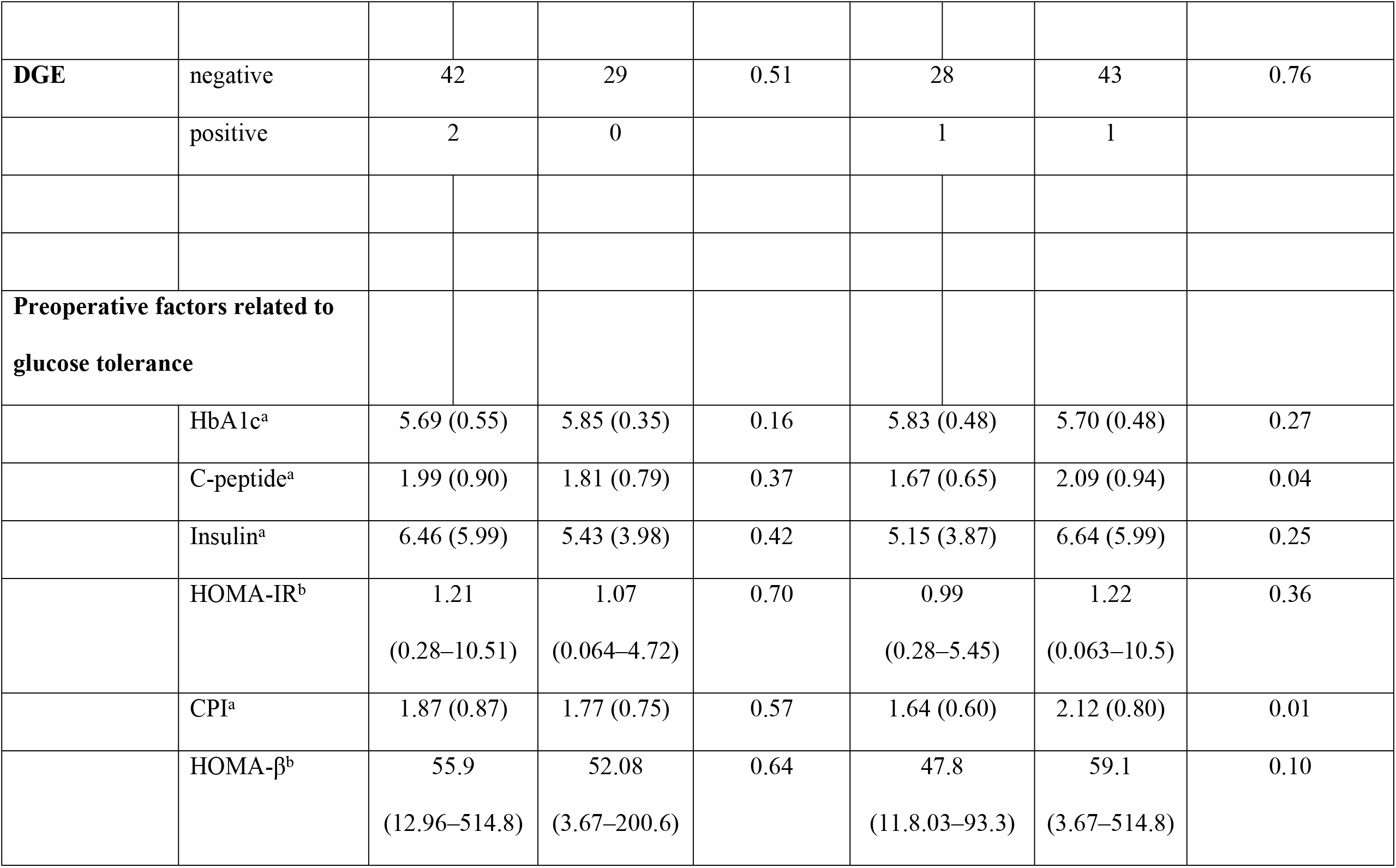
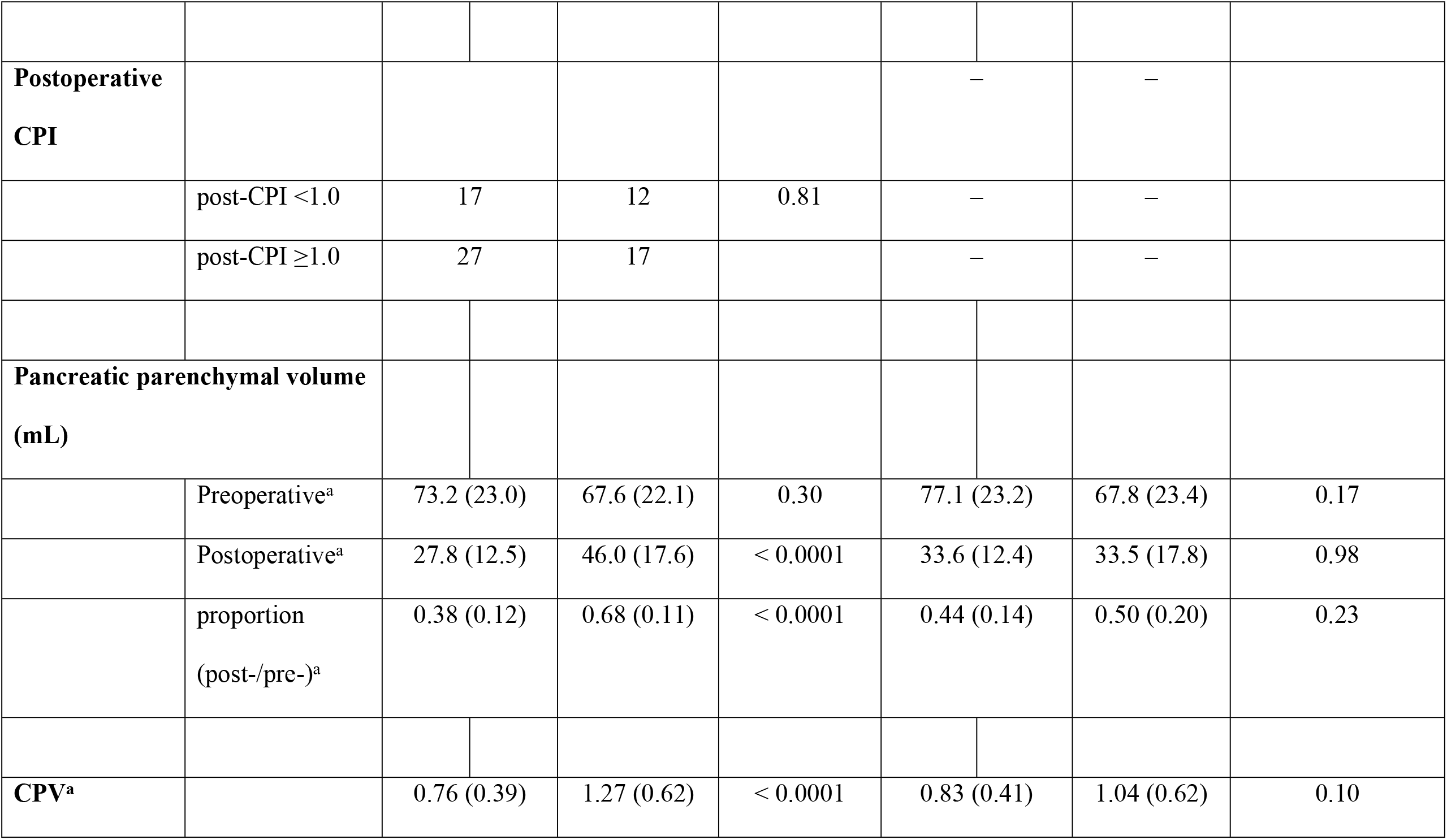
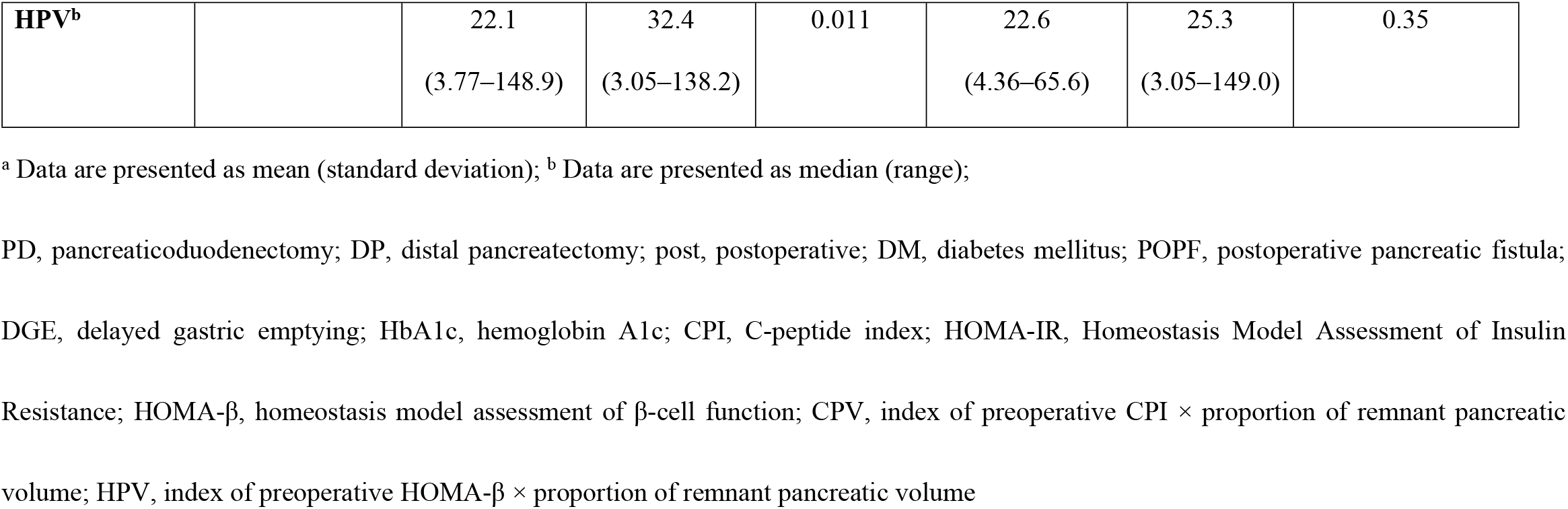
Comparison of Factors According to Surgical Procedure and Event Occurrence (Postoperative CPI <1.0) in Preoperative Non-diabetic Patients.

The changes in pre-and-postoperative CPI are shown in Fig 2. The CPI decreased postoperatively in 85% of the preoperative non-diabetic patients; post-CPI was significantly lower than pre-CPI in patients with these two operative procedures (Fig 2A, PD: p<0.001) (Fig 2B, DP: p<0.001).

**Fig 2.**
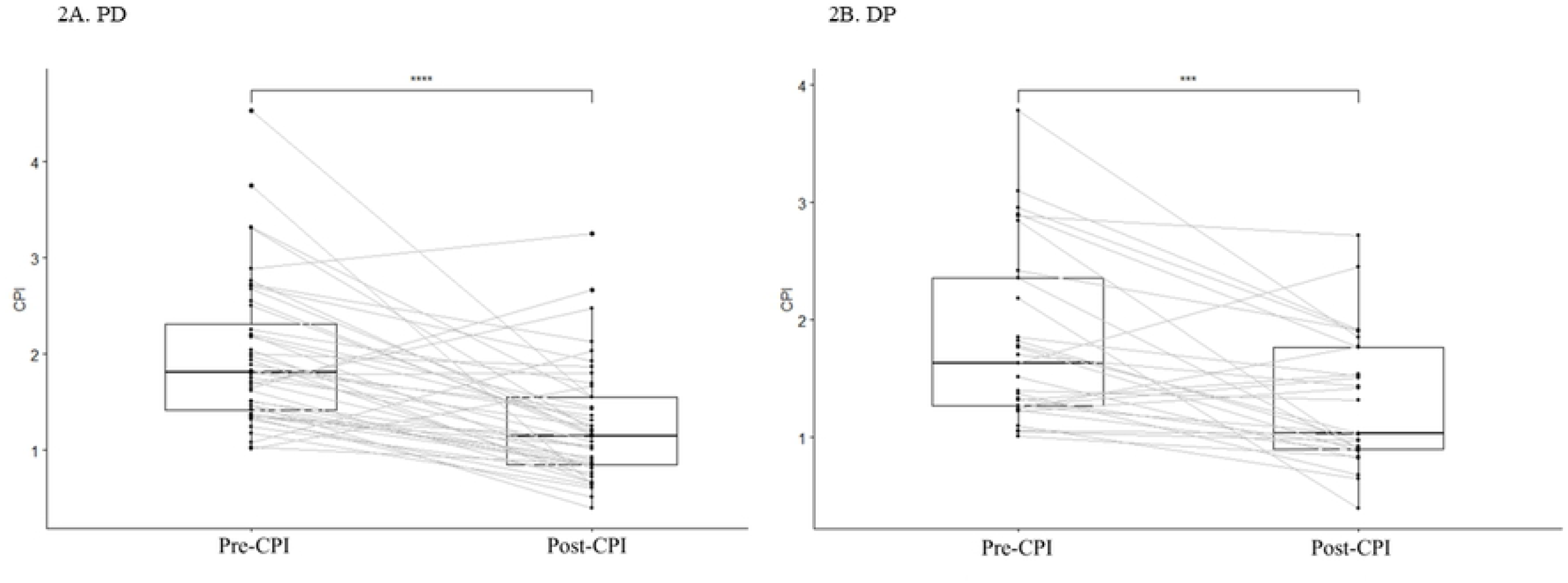
Changes in Pre- and Postoperative C-peptide Index (CPI) in Preoperative Non-Diabetic Patients for Each Surgical Procedure. A, PD; B, DP. PD, pancreaticoduodenectomy; DP, distal pancreatectomy; Pre, preoperative; Post, postoperative.

In preoperative non-diabetic patients who underwent PD, the median preoperative and postoperative CPIs were 1.81 (range: 1.01–4.54) and 1.15 (range: 0.40–3.26), respectively (Fig 2A). The Wilcoxon signed-rank sum test for the two corresponding groups (pre- and post-operative CPI) showed a significant difference with p=1.6×10^−6^ (Fig 2A). In preoperative non-diabetic patients who underwent DP, the median preoperative and postoperative CPIs were 1.63 (range: 1.01–3.79) and 1.03 (range: 0.40–2.72), respectively (Fig 2B). A significant difference was noted among the two corresponding groups (pre- and post-operative CPI), with p=2.7×10^−4^ (Fig 2B).

To assess the ability of each factor to predict the occurrence of post-CPI <1.0, we used the ROC-AUC. Preoperative CPI (pre-CPI) was the best predictor of the occurrence of post-CPI <1.0 in PD patients (Table 3, AUC: 0.75, 95% confidence interval (CI): 0.59–0.90). Similarly, pre-BMI was the most useful predictor in DP patients (Table 3; AUC: 0.68, 95% CI: 0.51–0.85). Consequently, logistic regression analysis after adjustment for age and sex was performed for pre-CPI in PD patients and pre-BMI in DP patients. Pre-CPI for PD patients was significantly associated with the event occurrence (post-CPI <1.0) in the adjusted multivariate analysis (Table 4). While in DP, pre-BMI could not be associated with an independent predictor of the event occurrence.

**Table 3.** Evaluation of the Predictive Ability of the Event Occurrence (Postoperative CPI <1.0) According to the Area Under the Receiver Operating Characteristic Curve.

| Covariate | PD | DP |
| --- | --- | --- |
| pre-BMI | 0.68 | 0.65 |
| pre-HbA1c | 0.61 | 0.53 |
| pre-CPI | 0.75 | 0.63 |
| CPV | 0.65 | 0.55 |
| pre-HOMA-β | 0.69 | 0.47 |
| HPV | 0.67 | 0.44 |
| pre-C-peptide | 0.67 | 0.59 |
| pre-insulin | 0.63 | 0.45 |
| **pre-HOMA-IR** | 0.63 | 0.47 |
| **PRPV** | 0.54 | 0.54 |

ROC, receiver operator characteristics; AUC, area under the curve; pre-, preoperative; BMI, body mass index; HbA1c, hemoglobin A1c; CPI, C-peptide index; CPV, index of preoperative; CPI × proportion of remnant pancreatic volume; CI, confidence interval; PD, pancreaticoduodenectomy; DP, distal pancreatectomy; HOMA-β , homeostasis model assessment of β-cell function; HPV, index of preoperative HOMA- $\beta \times$ proportion of remnant pancreatic volume; HOMA-IR, Homeostasis Model Assessment of Insulin Resistance; PRPV, proportion of remnant pancreatic volume

**Table 4.** Multivariate Logistic Regression Analysis of Predictive Factors for the Event Occurrence (Postoperative CPI <1.0) in Preoperative Non-diabetic Patients.

| **Covariate** | **Category** | **Odd ratio** | **95% CI** | **p value** |
| --- | --- | --- | --- | --- |
| **PD** |  |  |  |  |
| Age (years) | <68 | 1.11 | 0.29–4.29 | 0.87 |
| Sex | Female | 2.54 | 0.66–9.72 | 0.17 |
| *pre-CPI* |  | 0.27 | 0.081–0.94 | 0.038 |
| **DP** |  |  |  |  |
| Age (years) | < 68 | 1.35 | 0.26–6.93 | 0.720 |
| Sex | Male | 1.92 | 0.33–11.2 | 0.46 |
| *pre-BMI* |  | 1.31 | 0.55–1.03 | 0.082 |

CPI, C-peptide index; CI, confidence interval; pre, preoperative; PD, pancreaticoduodenectomy; DP, distal pancreatectomy

With regard to predicting the event occurrence (postoperative CPI <1.0) for each factor using the ROC-AUC, pre-CPI was the most useful predictor in PD (AUC: 0.750, 95% CI: 0.59–0.90) and pre-BMI was the most useful predictor in DP (AUC: 0.68, 95% CI: 0.51–0.85).

Logistic regression analysis, after adjustment for age and sex, was performed for pre-CPI for PD patients and pre-BMI for DP patients. Pre-CPI for PD patients was significantly associated with the event occurrence (post-CPI <1.0) in the adjusted multivariate analysis. While in DP, pre-BMI could not be associated with an independent predictor of the event occurrence. The stratified cross-validation results are shown in Table 5. As a predictive marker for the post-CPI <1.0 event occurrence, pre-CPI in PD demonstrated reasonable generalization performance.

**Table 5.** Mean Value of ROC-AUC after 10 Times Stratified Four-fold Cross-validation in Preoperative Non-diabetic Patients who Underwent PD.

| Covariate | Train data (SE) | Test data (SE) |
| --- | --- | --- |
| **pre-CPI** | 0.74 (0.047) | 0.83 (0.16) |

ROC, receiver operating characteristics; AUC, area under the curve; PD,
pancreaticoduodenectomy; SE, standard error; pre-, preoperative; CPI, C-peptide index

The cutoff value for predicting post-CPI <1.0 was 1.63 (sensitivity: 0.81, specificity: 0.71) for pre-CPI in preoperative non-diabetic patients who underwent PD (Table 6).

**Table 6.** Cutoff Value of the Predictive Marker for the Event Occurrence (Postoperative CPI <1.0) in Patients with Preoperative Non-diabetes who Underwent PD.

| Covariate | Cutoff value | Sensitivity | Specificity | PPV | NPV |
| --- | --- | --- | --- | --- | --- |
| **PD** |  |  |  |  |  |
| pre-CPI | 1.63 | 0.81 | 0.71 | 0.81 | 0.71 |

CPI, C-peptide index; PPV, positive predictive value; NPV, negative predictive value; PD, pancreaticoduodenectomy

Finally, pre- and postoperative glucose tolerance and postoperative insulin induction rate were compared between preoperative non-diabetic patients and patients on preoperative diabetic medication. Preoperative non-diabetic patients had significantly better preoperative HbA1c (Table 7, mean [SD]: 5.75 [0.48] vs. 6.60 [0.87], p<0.0001) and preoperative CPI (Table 7, 1.92 [0.76] vs. 1.42 [0.99], p<0.0001). Although the postoperative CPI and event occurrence (post-CPI <1.0) rates were not significantly different (Table 7, 1.27 [0.58] vs. 1.18 [0.90], p=0.60) and (Table 7, 39.7% vs. 56.2%, p=0.23), respectively, the postoperative insulin induction rate was significantly lower in the preoperative non-diabetic patients (Table 7, 1.4% vs. 37.5%, p<0.0001).

**Table 7.**
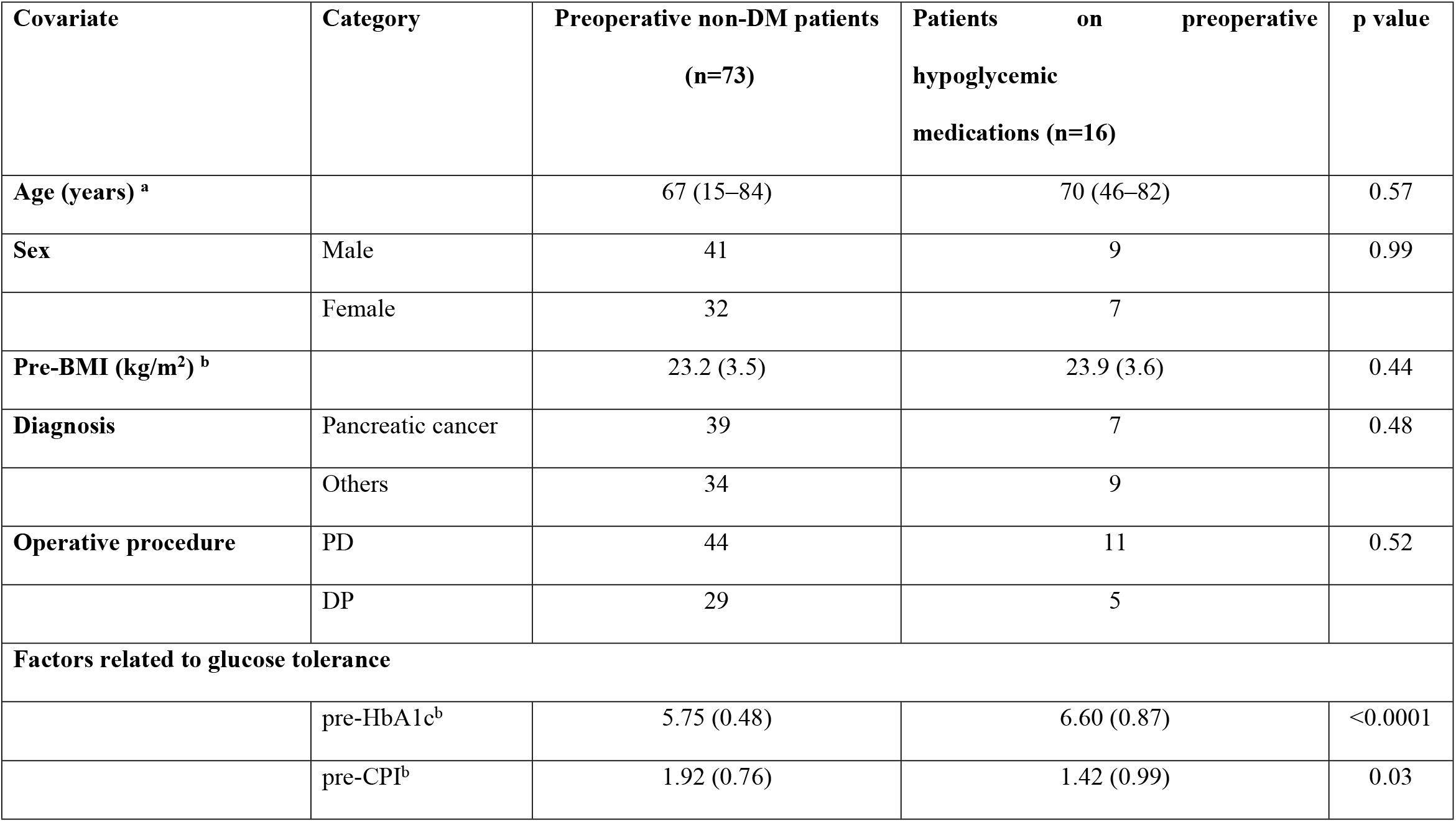
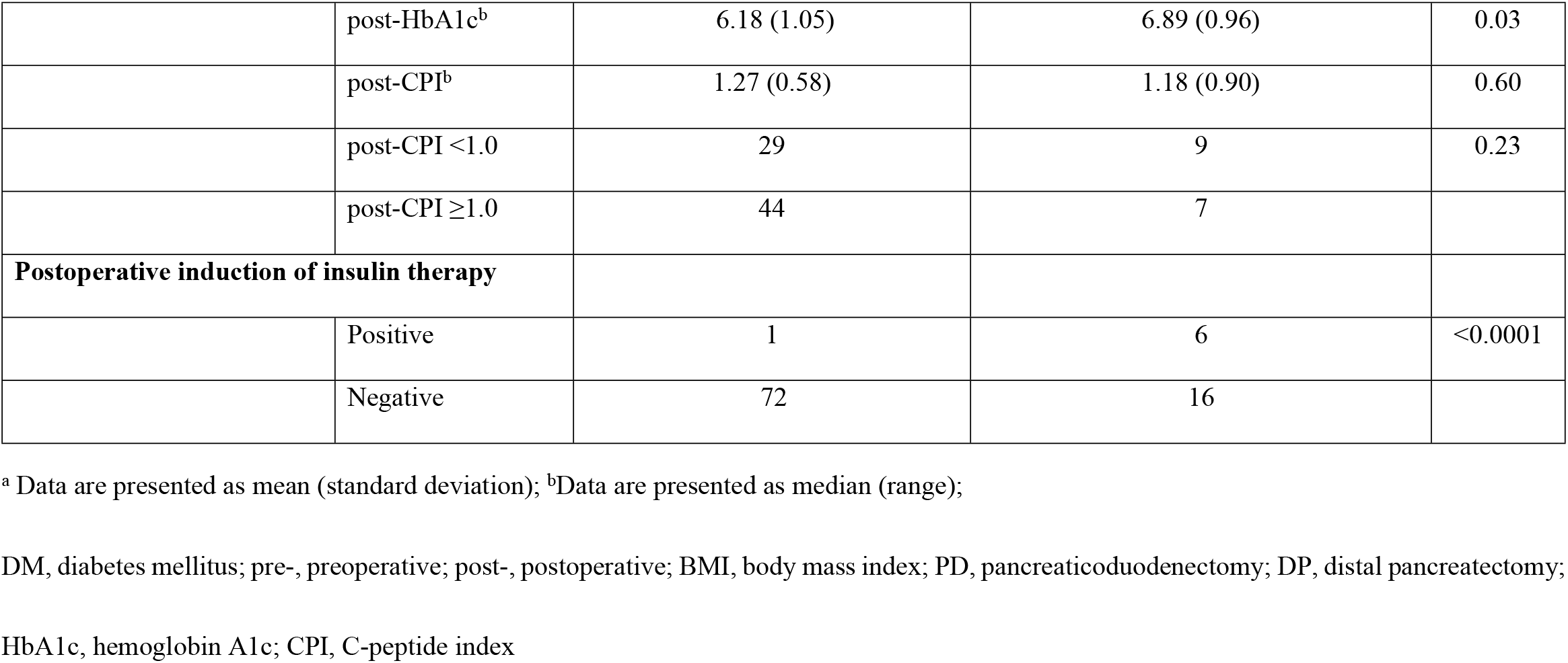
Comparison of Glucose Tolerance-related Factors Between Preoperative Non-diabetic Patients and Patients on Medical Therapy.

| **Covariate** | **Category** | **Preoperative non-DM patients**<br>**(n=73)** | **Patients on preoperative**<br>**hypoglycemic**<br>**medications (n=16)** | **p value** |
| --- | --- | --- | --- | --- |
| **Age (years) <sup>a</sup>** |  | 67 (15–84) | 70 (46–82) | 0.57 |
| **Sex** | Male | 41 | 9 | 0.99 |
|  | Female | 32 | 7 |  |
| **Pre-BMI (kg/m<sup>2</sup>) <sup>b</sup>** |  | 23.2 (3.5) | 23.9 (3.6) | 0.44 |
| **Diagnosis** | Pancreatic cancer | 39 | 7 | 0.48 |
|  | Others | 34 | 9 |  |
| **Operative procedure** | PD | 44 | 11 | 0.52 |
|  | DP | 29 | 5 |  |
| **Factors related to glucose tolerance** |  |  |  |  |
|  | pre-HbA1c<sup>b</sup> | 5.75 (0.48) | 6.60 (0.87) | <0.0001 |
|  | pre-CPI<sup>b</sup> | 1.92 (0.76) | 1.42 (0.99) | 0.03 |
|  | post-HbA1c<sup>b</sup> | 6.18 (1.05) | 6.89 (0.96) | 0.03 |
|  | post-CPI<sup>b</sup> | 1.27 (0.58) | 1.18 (0.90) | 0.60 |
|  | post-CPI <1.0 | 29 | 9 | 0.23 |
|  | post-CPI ≥1.0 | 44 | 7 |  |
| **Postoperative induction of insulin therapy** |  |  |  |  |
|  | Positive | 1 | 6 | <0.0001 |
|  | Negative | 72 | 16 |  |

<sup>a</sup> Data are presented as mean (standard deviation); <sup>b</sup>Data are presented as median (range);
DM, diabetes mellitus; pre-, preoperative; post-, postoperative; BMI, body mass index; PD, pancreaticoduodenectomy; DP, distal pancreatectomy;
HbA1c, hemoglobin A1c; CPI, C-peptide index

## Discussion

We used CPI as an index of BISC, and a postoperative CPI <1.0 was defined as a decrease in BISC that marked the occurrence of the event of interest. Postoperative CPI was significantly lower than preoperative CPI in approximately 85% of the patients, regardless of surgical procedure. The proportion of remnant pancreas volume by surgical procedure was significantly greater in DP than in PD, but there were no significant differences in the event rates. We identified preoperative CPI in PD patients as a predictor of post-CPI <1.0, while no significant factors were observed for DP patients. Preoperative non-diabetic patients had a significantly lower rate of insulin therapy induction after pancreatectomy than preoperative medically treated patients. In a comparison between preoperative non-diabetic patients and patients on diabetic medication, there were no significant differences in the postoperative CPI and event occurrence rates. However, the postoperative insulin induction rates were significantly lower in preoperative non-diabetic patients.

A previous report suggested preoperative 24-hour urinary C-peptide excretion measurement and multiplication of this value by the predicted pancreatic volume as a predictor of postoperative endocrine function [11]. However, 24-hour urine storage is a complicated and burdensome test for patients and is considered less versatile. However, CPI is a simple indicator that can be measured using blood tests and is considered more versatile.

The results of this study suggest that in terms of predictors of postoperative decreased BISC, the factors that are significant in PD are not significant in DP, suggesting that the predictors differ by surgical procedure. This difference might be due to the heterogeneous distribution of the islets of Langerhans in the pancreas. Generally, the islets of Langerhans are largely distributed in the pancreatic tail [4–6]. In PD, the pancreatic head, which has a large volume, is removed, while the pancreatic tail, which is thought to contain many islets of Langerhans, is preserved. Therefore, it was suggested that BISC after PD strongly reflects the index of preoperative BISC, which is represented by preoperative CPI. Since the residual pancreatic volume was large in DP cases, we evaluated the index considering the PRPV but did not obtain significant results. In a previous report, PRPV was not a risk factor for NODM after DP, and obese patients were significantly more likely to develop NODM [19]. Preoperative BMI was a candidate predictor of event occurrence in DP patients of this study, but it was not significant on multivariate analysis.

Regulation of insulin secretion is through stimulation by hyperglycemia or impact of various endocrine hormones. Incretin is a general term for gastrointestinal-derived hormones that act on pancreatic beta-cells to promote insulin secretion [20,21]. Glucagon-like peptide-1 (GLP-1) and gastric inhibitory peptide (GIP) are known as incretins, and their mechanisms of enhancing incretin effects and inhibiting metabolism have been elucidated and clinically applied as therapeutic targets for DM [20,21]. Muscogiuri et al. showed that duodenal resection in PD resulted in an increase in GLP-1 concentration, which was expected to increase insulin secretory capacity, but in fact, they reported a decrease in insulin secretory capacity [22]. This decrease is thought to be the result of the suppression of insulin secretion by increased glucagon levels [22]. Although we did not measure glucagon or incretin levels before and after surgery in this study, we believe that it is necessary to understand the behavior of hormones involved in glucose tolerance before and after surgery to elucidate the pathogenesis of changes in glucose tolerance due to pancreatectomy, and this is a topic for future research.

In the present study, most non-diabetic patients with post-CPI <1.0 did not progress to diabetes that would require insulin therapy postoperatively. It is speculated that not just a short-term decrease in BISC is involved in the development of NODM. Mezza et al. stated that even if there is forced removal of β-cells by pancreatectomy, the function of the remaining β-cells will not lead to insulin deficiency if there is no insulin resistance [23]. However, if there is insulin resistance at the time of pancreatectomy, the residual β-cells overload postoperatively, leading to insulin deficiency and NODM. In other words, the true determinant of the development of NODM is not forced removal of β-cells by pancreatectomy but the presence or absence of a pre-existing diabetic milieu, which may lead to type 2 DM, a condition accelerated by surgery [23]. Similar to the previous report, in this study, preoperative non-diabetic patients had a significantly lower rate of postoperative insulin introduction than patients on preoperative diabetic medication. These results suggest that the coexistence of pancreatectomy induced BISC reduction and insulin resistance, which is considered one of the pathophysiological features of type 2 DM, may promote postoperative insulin deficiency, resulting in the need for insulin therapy.

Although the etiology of insulin resistance is not fully understood, obesity has been identified as an acquired risk [23]. Patients who are expected to have decreased BISC after pancreatectomy may reduce the development of NODM by weight control based on appropriate diet and exercise therapy, with appropriate intervention through follow-ups being essential in the future.

This study has some limitation, including the single-center, retrospective design. Using the three explanatory variables of age, sex, and predictors of event occurrence in multivariate analysis to determine minimum sample size, a minimum of 30 cases of event occurrence were required per surgical procedure, and assuming an event occurrence rate of 40%, the total number of cases should be at least 75 each per surgical procedure. Since the overall numbers of cases and events were limited in this study, further analysis with a larger number of cases is crucial. Furthermore, medium-to long-term follow-up is necessary to clarify how early postoperative decline in BISC changes over the medium-to long term and whether it leads to the development of NODM.

## Conclusion

Preoperative CPI was determined to be the most useful factor for predicting a decrease in BISC after pancreatectomy in patients who underwent PD. Predicting postoperative BISC decline and simultaneously confirming the presence of insulin resistance could be useful in predicting the development of NODM after pancreatectomy.

## Data Availability

Data are available from the Ethics Committee (contact via rhe responsible author) for researchers who meet the criteria for access to confidential data.

## Acknowledgments

The authors acknowledge the support of all medical and surgical staff who cared for the patients in this study.

